# Mapping the Spatial and Single-Cell Architecture of Cutaneous Squamous Cell Carcinoma Progression

**DOI:** 10.64898/2026.11.21.26368487

**Authors:** Kristin Peters, Felicitas Geserick, Lena Kohne, Anna Staffeld, Aenne Foth, Rüdiger Panzer, Lars Boeckmann, Steffen Emmert, Sander Bekeschus, Susann Groth, Matthias Sendler, Torben Redmer, Josefine Radke

**Affiliations:** Institute of Pathology, University Medicine Greifswald, Greifswald, Germany; Institute for Molecular Genomics and Core Unit Genomics (CUG), University Medicine Greifswald, Greifswald, Germany; Department of Dermatology, Venereology and Allergology, Rostock University Medical Center, Rostock, Germany; ZIK plasmatis, Leibniz Institute for Plasma Science and Technology (INP), Greifswald, Germany; Department of Internal Medicine A, University Medicine Greifswald, Greifswald, Germany; Institute of Pathology, Unit of Laboratory Animal Pathology, University of Veterinary Medicine Vienna, Austria; Comprehensive Cancer Center Mecklenburg-Vorpommern (CCC-MV), c/o Biotechnikum Greifswald, Greifswald, Germany

**Author notes:** These authors contributed equally as first authors (Kristin Peters, Felicitas Geserick). These authors contributed equally as senior/last authors (Torben Redmer, Josefine Radke).

## Abstract

Cutaneous squamous cell carcinoma (cSCC) represents approximately 20% of all skin cancers and is rising in global incidence. Although cSCC is known to progress from chronically sun-damaged keratinocytes via premalignant actinic keratosis (AK), the underlying spatial and molecular dynamics remain poorly defined. Here, we applied Xenium single-cell spatial transcriptomics to whole-skin sections from AK (n = 2) and cSCC (n = 2) patients to map key transitions in cSCC development. We resolved four distinct cSCC tumor cell subpopulations—epithelial, secretory, invasive, and stressed—whose transcriptional profiles diverge from bulk cSCC as well as normal and activated keratinocytes. Notably, we identified a MET^high^ tumor subpopulation, potentially linked to aggressive disease progression and resistance to immune checkpoint inhibition.

## Results and Discussion

Accounting for roughly 20% of skin cancers, cutaneous squamous cell carcinoma (cSCC) is the second most common non-melanoma skin cancer and shows a rising global incidence^1,2^. The tumor develops gradually through a transformation of keratinocytes, which initially leads to actinic keratoses (AK) - squamous cell carcinomas in situ precursor lesions that form on chronically sun-damaged skin through mechanisms that are not yet fully understood. In order to provide insights into the different steps of cSCC development, we performed Xenium-based single cell-resolved spatial transcriptomics (ST) profiling of whole skin sections of patients with AK (n=2; AK_I, AK_II) and cSCC (n=2; cSCC_1, cSCC_2) (Figure 1a, H&E and Figure S1a left, Table S1). Tissue sections were hybridized and scanned (Figure 1a), yielding 23 distinct clusters via marker-based profiling across all merged sections (Figure S1a, right). We have identified three functional subpopulations of cSCC tumor cells (TC)— epithelial, secretory, invasive, and stressed cells—whose molecular profiles differ significantly from those of general cSCCs as well as normal and activated keratinocytes (Figure 1a, center; Table S2). Crucially, we detected a MET^high^ cSCC population, a phenotype known to drive aggressive progression and confer resistance to immune checkpoint inhibition^3^. Expression profiling of skin samples revealed unique, stage-specific gene signatures (Figure S1b). Using the ESTIMATE algorithm to infer immune cell infiltration, we spatially mapped immune-enriched areas across individual sections, revealing distinct immune “hotspots” localized to highly inflamed sites (Figure 1a, right; Figure S1c). The tumor microenvironment (TME) likely associated with keratinocyte transformation comprised T cells, B cells, plasma cells, dendritic cells (DCs), tumor-associated macrophages (TAMs), cancer-associated fibroblasts (CAFs), and resident skin cells such as hair follicle stem cells (HFSCs), mast cells, and Langerhans cells (LCs) (right image in Figure 1a, Figure S1d, and Table S2). The spatial patterning of these tumor and TME cells likely shaped the ecosystems of early- and late-stage transformed keratinocytes in AKs and cSCC. When examining the individual cellular subfractions, we observed that the skin samples differed primarily in the proportions of these subfractions, but not in their composition (Supplementary material, Figure S1e). cSCC cells exhibited a pronounced loss of cellular polarity and disruption of the characteristic vertical alignment of keratinocytes (Figure S2a). Molecularly, differential expression (DE) analysis revealed cell type-specific gene signatures distinguishing both normal and activated keratinocytes (I) and normal keratinocytes and cSCC cells (II) (Table S3). DE analysis validated established markers lost during the cellular transition of healthy keratinocytes into AK, such as NOTCH1 and the major structural proteins of the spinous layer, KRT1 and KRT10^4^. The marker-based subclassification of MET^high^ cSCCs revealed a potentially highly invasive tumor subset, featuring high expression of TP63 and TNC (Figures 1b (IV) and S2b), probably driving tumor progression in concert with MET. MET and TNC expression was not observed in AKs and normal skin (Figure S2b) and likely serves as a specific marker of progressive cSCC. Spatial mapping of a core MET gene signature revealed the highest MET scores in cSCC samples (Figure S2c). CEACAM6 (p = 4.59 × 10^−13^, Wilcoxon rank-sum) and MET (p = 2.99 × 10^−2^, Wilcoxon rank-sum) significantly distinguished cSCC and MET^high^ cSCC (Figure S2d). Spatial analysis revealed distinct compartmentalization, with immune cell infiltration largely separated from tumor regions (Figure 1c). While T cells, B cells, and tumor-associated macrophages (TAMs) were sparsely dispersed throughout tumor areas, they primarily aggregated into dense immune clusters. Cell lineages were confirmed via immunostaining for pan-cytokeratin (tumor cells), CD3 (T cells), CD20 (B cells), and CD68 (macrophages) (Figure 1c, right). Multiplex immunofluorescence (MACSima) showed that B cell clusters contained highly proliferative (Ki67^+^) centers devoid of activated (HLA-DR^+^) T cells^5^ (Figure 1d (I), left). In contrast, T cell clusters contained partially activated CD4^+^ and CD8A^+^ T cells (Figure 1d (I), center). Single-cell-resolved spatial transcriptomics further identified dendritic cells (DCs), NK cells, and TAMs within T cell clusters, highlighting a CLEC12A^+^ DC/TAM subset associated with immune suppression^6^ (Figure S3a). With respect to immune topologies, we found that T cell clusters expressed pan-T cell (CD3, CD4), cytotoxic T cell (CD8A), naïve B cell (CD79A), and plasma cell (IGKC) markers, contrasting infiltrated tumor-adjacent areas that lacked both CD8A^+^ T cells and B cells (Figure 1d (I), right; Figure 1d (II)). In summary, our study provides single-cell spatial insights into the architecture of cutaneous squamous cell cancer progression from AK to cSCC, suggesting that immune cell recruitment coincides with early keratinocyte activation (AK), whereas immune cell exclusion accompanies disease progression to cSCC.

**Figure 1:**
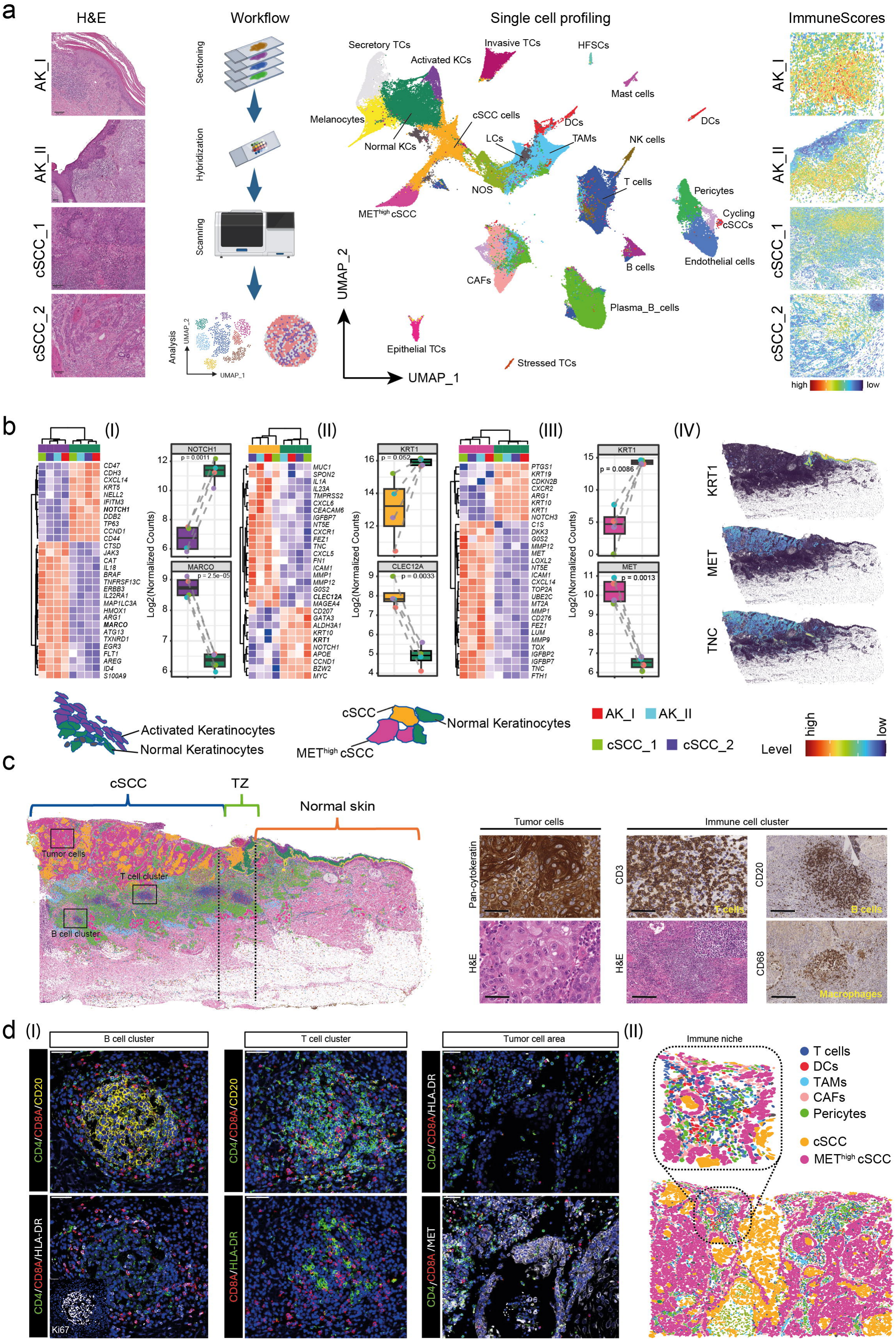
Single cell spatial profiling uncovers cellular subsets of cutaneous squamous cell carcinoma. **a.)** Hematoxylin and eosin (H&E) staining of representative areas of skin samples showing AK lesions (actinic keratosis, AK_I, AK_II) or presence of invasive cSCC (cSCC_1, cSCC_2). The workflow illustrates the different analysis steps: Sample sectioning, hybridization of sections with probes defined by the panels, scanning of hybridized tissue samples, collection of count data and spatial information, and subsequent analysis. Single sample profiling and UMAP presentation revealed marker-defined cellular subsets of cSCC, besides immune cells T cells, B cells, NK cells, Plasma B cells, DCs, Langerhans cells (LCs) and skin-resident Mast cells, hair follicle stem cells (HFSCs), normal keratinocytes (KCs), as well as cancer-associated cells, tumor-associated macrophages (TAMs), cancer-associated fibroblasts (CAFs), and activated KCs. Endothelial cells and pericytes account for both normal skin and skin with neoplasia (center). The spatial mapping of immune scores indicates areas of immune cell infiltration (right panels). **b.)** Heat maps showing differentially expressed genes of normal vs. AK (I), normal KCs vs. cSCC (II), and normal KCs vs. METhigh cSCC (III) across all tissue sections (tissue samples and cell types are color-coded). The spatial mapping of KRT1, MET, and TNC expression illustrates spatially distinct areas of gene expression (color-coded). **c.)** Spatial map and H&E image of sample cSCC_1 indicating different phenotypical states of cSCC cells and an increase in immune cell infiltration accompanying progression. Immunohistochemistry of selected areas (boxes) indicates the cellular origins: pan-cytokeratin (Tumor cells), T cells (CD3), B cells (CD20), and macrophages/TAMs (CD68). d.) Multiplex immunofluorescence analysis (MACSima) of selected areas showing a B cell cluster (left), T cell cluster (center), and tumor cells (right) (I). Schematic representation of an immune niche.

## Data Availability

The supplementary figures, tables, and raw datasets generated in this study are publicly available in Zenodo at https://doi.org/10.5281/zenodo.24167237.

## Authors’ contributions

K.P., F.G., M.S., J.R., and T.R. conceived and designed the study, performed experiments, collected data, analyzed the data, and drafted and wrote the manuscript. L.K., A.S., A.F., R.P., and L.B. performed experiments and provided essential resources. S.E., S.B., and S.G. contributed substantially to the conceptual development and critical revision of the manuscript. All authors critically revised the manuscript for intellectual content and approved the final version for submission.

## Acknowledgements

We would like to thank all collaborators, technical staff, and laboratory personnel who contributed to this study. We are especially grateful to Andreas Wendt and Dana Ristau for their support with sample preparation, immunohistochemistry, and technical assistance.

