## Supplementary material for "Mapping the Spatial and Single-Cell Architecture of Cutaneous Squamous Cell Carcinoma Progression"

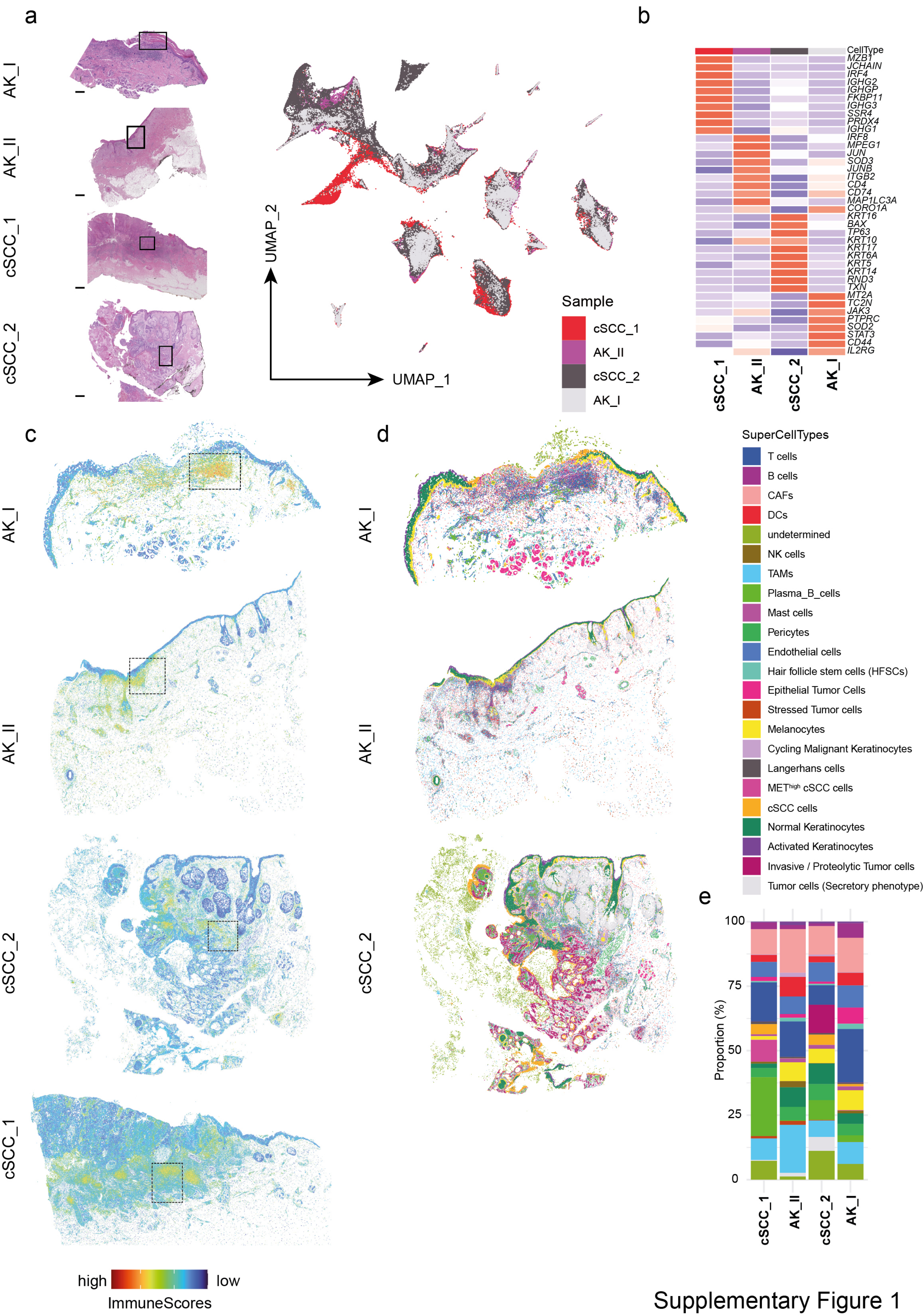


Supplementary Figure 1

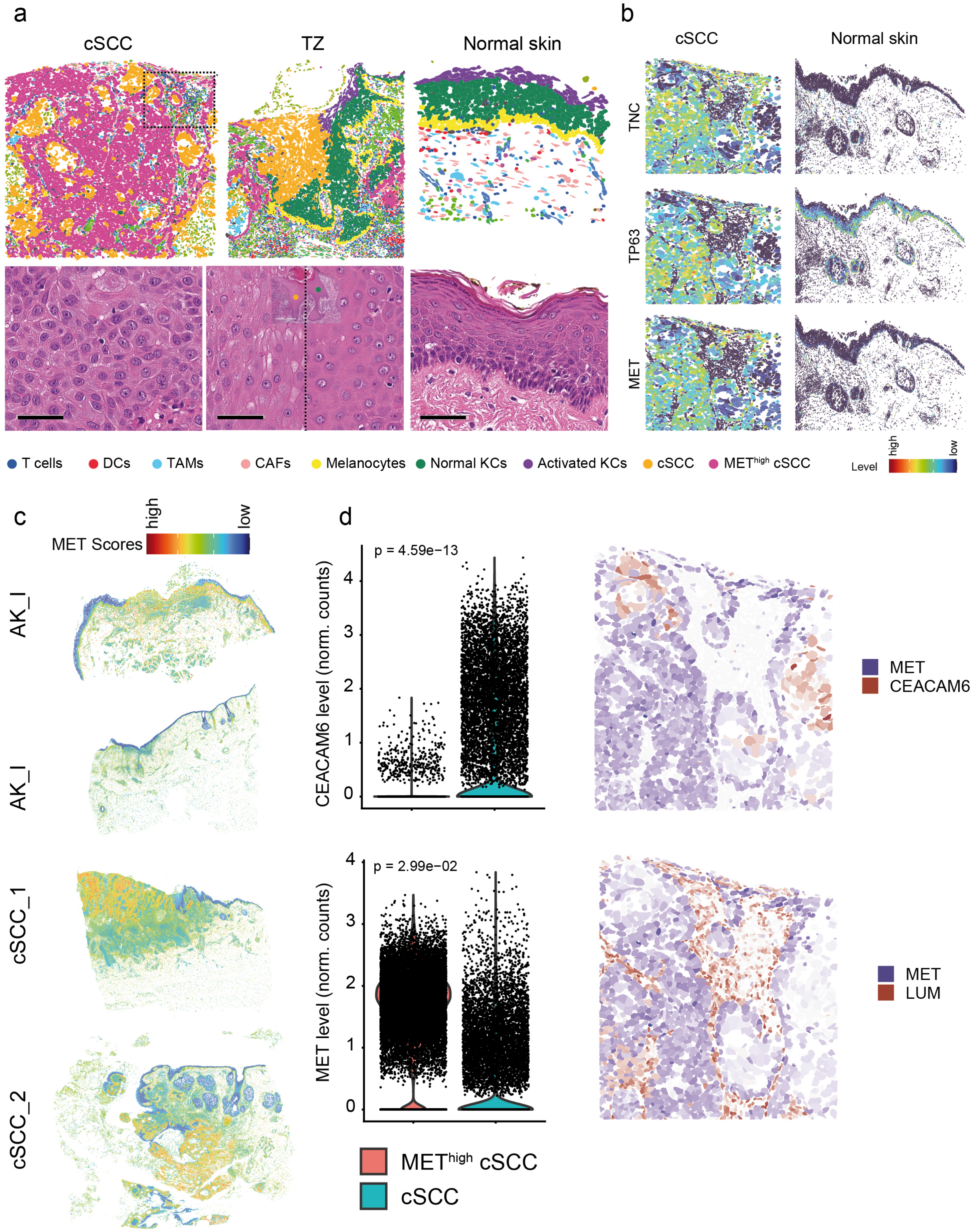


Supplementary figure 2

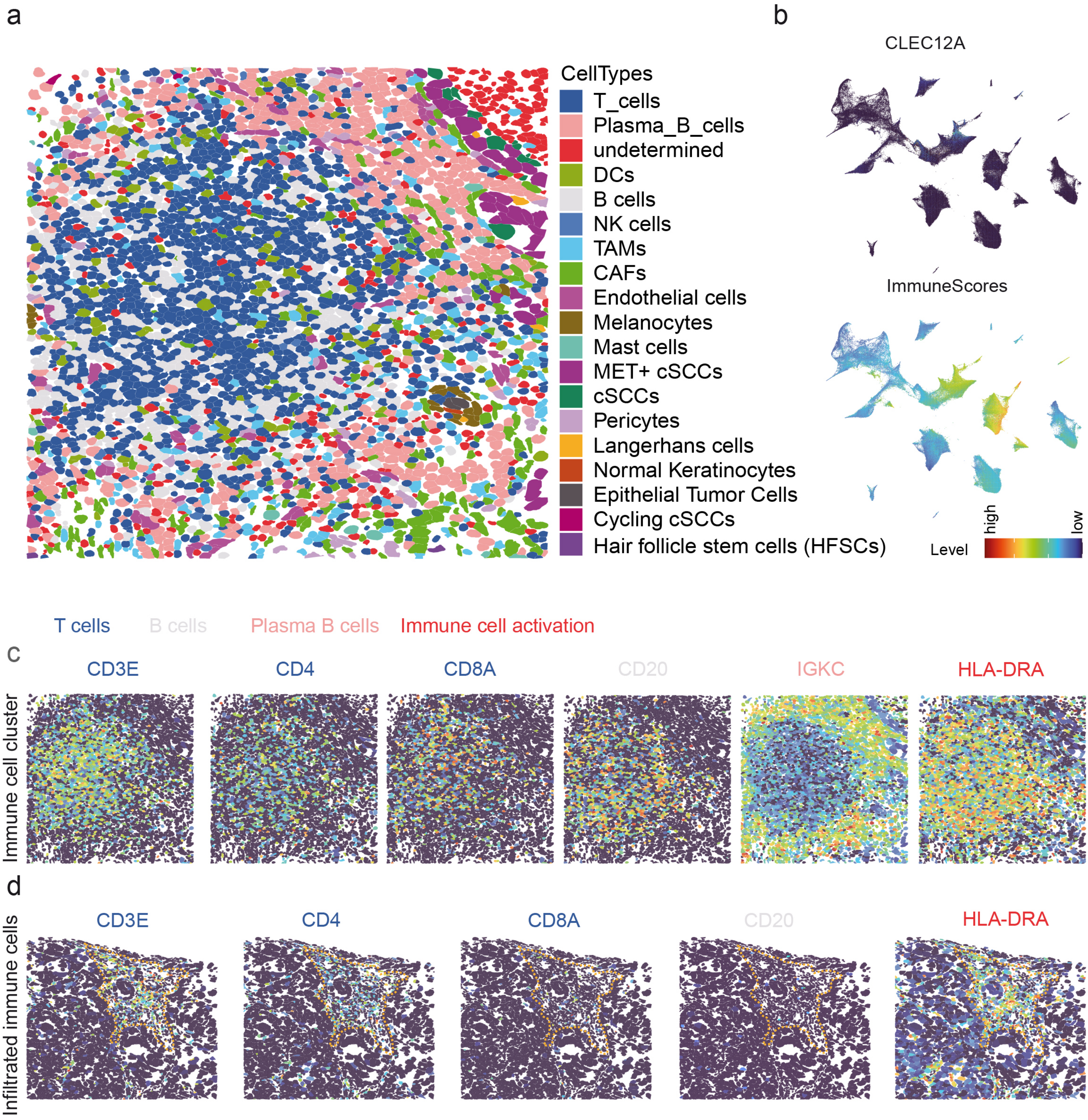


Supplementary figure 3

### Mapping the Spatial and Single-Cell Architecture of Skin Cancer Progression

#### Supplementary Material and Methods

##### Patient samples

Formalin-fixed and paraffin embedded (FFPE) archived skin samples of patients diagnosed with actinic keratosis or squamous cell carcinoma (Table S1) were collected at the Department of pathology, University Medicine Greifswald (UMG) and stored at room temperature. All samples have been examined by a trained pathologist.

##### Spatial transcriptomics analysis and Data processing

Xenium in situ analysis (10X Genomics) was performed at the Spatial & Functional Screening Core Facility, Heinrich Heine University Düsseldorf. Skin sections were processed by Dr. Yi-Nan, according to manufacturer's guidelines (CG000578, Rev D; CG000760, Rev A). Briefly, 5 µm sections underwent sequential deparaffinization and permeabilization, followed by overnight hybridization with a custom probe panel (100 genes) which was combined with the pre-designed immuno-oncology panel (10X Genomics, 380 genes) at 50°C, rolling circle amplification, and slide-read out in the Xenium Analyzer. After run completion, H&E staining was performed following Xenium guidelines (CG000613, B). Data were processed using the Xenium Onboard Analysis pipeline, Xenium output files were subsequently analyzed in R and RStudio (Version 2026.09.0+174 "Autumn Hawkbit" as described previously. Briefly, spatial analysis of the Xenium readouts were analyzed using the VoltRon (version 0.2.2) R package[1]. Individual sections (n=4) were grouped into patient/donor of origin prior to downstream analysis. Cells with transcript counts less than 5 were removed prior to clustering. Seurat (version 5) package[2] was used to perform downstream unbiased clustering of Xenium cells. Subsequently, dimensionality reduction is performed using PCA and UMAP, followed by clustering with shared-nearest-neighbor graphs (the "FindNeighbors" and "FindClusters" functions) using parameters specific to each sample. UMAP of Xenium cells across all sections were calculated using Harmony-based[3] integrated PCA embeddings. Marker analysis was performed to detect differentially expressed (DE) genes specific to each cell cluster (FindAllMarkers function). We used the AddModuleScore function from Seurat to calculate the different scores (ImmuneScore, METScore, cSCCScore) using identified markers or a predetermined set of genes (ESTIMATE). To detect significant changes of gene expression between defined cellular subsets across sections we aggregated the count of annotated tumor cells and cells of the TME regarding the cellular subtype and performed DE analysis with the DESeq2 R package[4].

**Immunohistochemistry**

Automated immunohistochemical staining of formalin-fixed, paraffin-embedded (FFPE) tissue sections (2 µm thick) was performed on a VENTANA Benchmark XT automated staining instrument (Ventana Medical Systems, Tucson, AZ) according to the manufacturer's instructions as previously described5. Immune cells were detected using primary antibodies for CD3 (DAKO, polyclonal, rabbit), CD20 (DAKO, clone LN26, mouse) and CD68 (DAKO, clone KP1, mouse). A pan-cytokeratin (AE1/3, DAKO, clone A1/A3 mouse) antibody was used for tumor cell detection. For detection, the ultraView Universal DAB Detection Kit (DAKO) was applied.

**MACSima-based multiplex imaging**

FFPE tissue sections (2 µm) were were prepared according to the manufacturer's protocol ("MACSima™ Imaging Cyclic Staining (MICS) Sample preparation protocol for FFPE tissue" by Miltenyi Biotec). After incubation at 60°C for one hour the sections were left to dry at room temperature overnight and processed for multiplex staining as preciously described[5]. The following directly fluorescently labeled primary antibodies (Miltenyi Biotec) were used for the detection of CD4 (diluted 1:50), CD8a (diluted 1:50), CD20 (diluted 1:50), HLA-DR (diluted 1:50), c-MET (diluted 1:50) and Ki67 (diluted 1:50). Following the MACSima run, H&E staining of sections was performed and scanned using the Sysmex Slide Scanner MIDI-II from 3D Histech.

#### Supplementary figure legends

**Figure S1: Skin samples show proportional differences but compositional congruence. a.)** H&E stained whole sections of skin samples, selected areas are indicated. Scale bars indicate 3 mm (left). UMAP presentation of single cells of combined, clustered and harmonized skin samples (right). **b.)** Top differentially expressed genes among skin samples, showing sample specificity. **c.)** Spatial mapping of immune scores. Areas of high immune scores (color coded) correspond to immune cell enriched sites. **d.)** Spatial maps of skin samples indicating the presence and location of cellular subsets as indicated. **e.)** Proportions of cellular subsets shown in (d).

**Figure S2: Expression of CEACAM6 and MET discriminate cSCC and MET$^{high}$ cSCC. a.)** Selected areas of sample cSCC_1 indicating the tumor area (cSCC), the transition zone (TZ), the crossing from normal skin into the area of keratinocyte transformation and normal skin. **b.)** Comparative mapping of expression of tenacinC (TNC), TP63 and MET to cSCC and normal skin, indicating enriched gene expression in the tumor cells. **c.)** Spatial mapping of the METScore to skin samples indicating areas of expression of MET and MET-associated genes which is highest in cSCC samples and low in AK samples. **d.)** Box plots (left) and spatial maps (right) depicting the levels and spatial location of CEACAM6 and MET across single cells in cSCC and MET$^{high}$ cSCC. Significance was determined by Wilcoxon test. The mapping of lumican (LUM) and MET indicates the stromal cell compartment and tumor cell compartment (right, bottom).

**Figure S3: Spatial and single cell resolution of a T cell cluster revealed cellular heterogeneity. a.)** Spatial map of a T cell enriched cell cluster indicating several cell types determining the cluster or surrounding the cluster. **b.)** UMAP presentation of single cells of skin samples showing levels of CLEC12A expression (top) and levels of immune scores (bottom).

**c.)** Spatial mapping of expression of CD3E, CD4, CD8A, CD20, IGKC and HLA-DR in single cells of the T cell cluster shown in (a). **d.)** Spatial mapping of expression of CD3E, CD4, CD8A, CD20 and HLA-DR in single tumor infiltrated immune cells. In b, c, d the expression level is color coded.

#### Supplementary table legends

**Table S1: Patients characteristics.** TNM stage, age, gender and the main histopathological findings are reported.

**Table S2: Marker-based classification of cellular subsets.** Every cellular subset is provided by a gene signature, expression level (log2FoldChange) and significance of expression of a certain cluster compared with all clusters.

**Table S3: Differentially expressed genes determining tumor cell subsets.** DESeq2 analysis was performed to determine the genes specifically determining normal KCs, activated KCs, cSCCs or $MET^{high}$ cSCCs
